# Refining COVID-19 retrospective diagnosis with continuous serological tests: a Bayesian mixture model

**DOI:** 10.1101/2023.09.15.23295603

**Authors:** Benjamin Glemain, Xavier de Lamballerie, Marie Zins, Gianluca Severi, Mathilde Touvier, Jean-François Deleuze, SAPRIS-SERO study group, Nathanaël Lapidus, Fabrice Carrat

## Abstract

COVID-19 serological tests with a “positive”, “intermediate” or “negative” result according to predefined thresholds cannot be directly interpreted as a probability of having been infected with SARS-CoV-2. Based on 81,797 continuous anti-spike tests collected in France after the first wave, a Bayesian mixture model was developed to provide a tailored infection probability for each participant. Depending on the serological value and the context (age and administrative region), a negative or a positive test could correspond to a probability of infection as high as 61.9% or as low as 68.0%, respectively. In infected individuals, the model estimated a proportion of “non-responders” of 14.5% (95% CI, 11.2-18.1%), corresponding to a sub-group of persons who exhibited a weaker serological response to SARS-CoV-2. This model allows for an individual interpretation of serological results as a probability of infection, depending on the context and without any notion of threshold.

## 1 Introduction

Provided with the result of a dichotomous diagnostic test (“positive” or “negative”), estimating the probability that a disease of interest is present is not straightforward. This post-test probability can be computed according to Bayes’ rule using estimates of the disease’s frequency (“prior probability”) and of the test’s sensitivity and specificity (“likelihood”), but complexity arises when modeling biological and epidemiological features of the pair disease-test and estimating uncertainty (confidence intervals) to correctly reflect the amount of information at hand.

In that sense, serosurveys that took place after the first wave of SARS-CoV-2 pandemic to quantify its extent were facilitated by convenient kinetic properties of biological response in an immunologically naive population [1]. Indeed, as symptoms can follow infection by a week (on average) and hospitalization follows symptoms by another week (if so), anti-spike immunoglobulin G (IgG) rises within a month after the infection and remain stable during several months (contrary to other immunoglobulins or to vaccine response) [2–7]. To summarise, in the summer of 2020, anti-spike IgG of infected persons had already risen but had not yet decreased, which in particular allowed for the use of cross-sectional methodologies in serosurveys. In France, several studies were based on a test produced by Euroimmun that measured the presence of IgG targeting the S1 domain of the SARS-CoV-2 spike protein [8–10]. Seroprevalence (the proportion of samples above the 1.1 cut-off value recommended by the manufacturer) was estimated to be about 5% in the whole country and 10% in Ile-de-France (Paris area). This 1.1 threshold (optical density ratio) was associated with a sensitivity of 91.4% (92.7% when excluding tests realized less than 14 days after symptoms onset) and a specificity of 98.6% [11].

At the population scale, deducing cumulative incidence (the proportion of persons having been infected since the beginning of the pandemic) from seroprevalence requires to take sensitivity and specificity into account, possibly through Bayesian methods (as a mean to preserve uncertainty) [12]. This process usually involves three samples: two of them include individuals of known infection status to estimate the test’s characteristics (sensitivity from infected individuals and specificity from uninfected ones), and the third sample includes individuals of unknown infection status (undetermined sample) from which cumulative incidence is estimated. Methods like post-stratification can be applied if this third sample does not correctly represent the target population (selection bias). A spectrum bias occurs when the sample of known infected is not representative of infected individuals in the population under study. This bias is often suspected, as symptomatic individuals are more likely to be detected and therefore recruited to study sensitivity, and are also more likely to have higher antibody levels [13–16]. In that case, a sensitivity parameter can be estimated in each infected sub-group (symptomatic or asymptomatic). Cumulative incidence will then be computed using the appropriate sensitivity parameter, according to the presence of symptoms in the unknown status sample, necessitating to record the presence of symptoms during data collection. Mixture models constitute an appealing solution in the case where such variables are not available in the data, as continuous distributions of serological results can be estimated for both groups (infected and uninfected) directly from the sample of undetermined infection status (hence not relying entirely on biased validation samples) [17, 18]. Briefly, a mixture model can be described in the present case as a weighted average of several continuous probability distributions (one distribution for each sub-population, the weights being the proportions of these sub-populations in the overall population). These models are however prone to identification issues, corresponding to situations where more than one tuple of parameters’ values are consistent with the data (this happens notably when one sub-population can “explain” the whole data mixture) [19]. As a result, MCMC convergence issues can occur due to several local maxima in the posterior distribution of parameters.

Finally, predictions at the individual scale (in the present case, retrodictions) should take advantage of covariates that influence cumulative incidence, test’s characteristics, as well as all the information contained in the test. Concretely, keeping the test continuous by the means of a mixture model has been shown to outperform cut-off-based methods (which dichotomize the test) in terms of accuracy [18].

This study aimed at refining COVID-19 retrospective diagnostic at the individual scale by applying a Bayesian mixture model to continuous serological data collected in France after the first wave of the pandemic, taking age and geographic area into account. Plus, the model was used to quantify a proportion of “non-responders” (an imperfectly-defined group of persons whose antibody levels do not increase, or increase only slightly, after a SARS-CoV-2 infection), which has been reported to lie between 5 and 24% depending on classification criteria and methodologies [20, 21].

## 2 Results

### 2.1 Participants

All samples were collected between May and November, 2020. Among the total cohort of 82 467 individual with one serological test, 319 had a positive RT-PCR test and constituted the sample with known infection (mean age of 52 years, 29% men, mean elapsed time between the RT-PCR and dried blood sampling of 100 days, with a minimum of 12 days and a maximum of 190 days). After excluding 351 samples of individuals with missing data on administrative region of residence, the sample of undetermined status individuals included the remaining 81 797 individuals (mean age 58 years, 35% men). No known uninfected group was available. The number of observations for each region and each age group is provided in Supplementary Tables 1 and 2. Corsica was not considered in the illustrative example of infection retrodiction below due to the low count of tests made in this region.

### 2.2 Cumulative incidence and infection-outcome rates

Cumulative incidence of COVID-19 among adults in metropolitan France after the first wave was 7.6% (95% CI, 7.3 to 7.9%), with a peak at 11.7% (95% CI, 11.0 to 12.4%) in Île-de-France (Paris area). Figure 1 features a map of metropolitan France showing the heterogeneity in cumulative incidence associated with location (exhaustive regional estimates are provided in Supplementary Table 3). IHR and IFR at the scale of metropolitan France were 2.6% (95% CI, 2.5 to 2.7%) and 0.8% (95% CI, 0.8 to 0.9%), respectively.

**Fig. 1.**
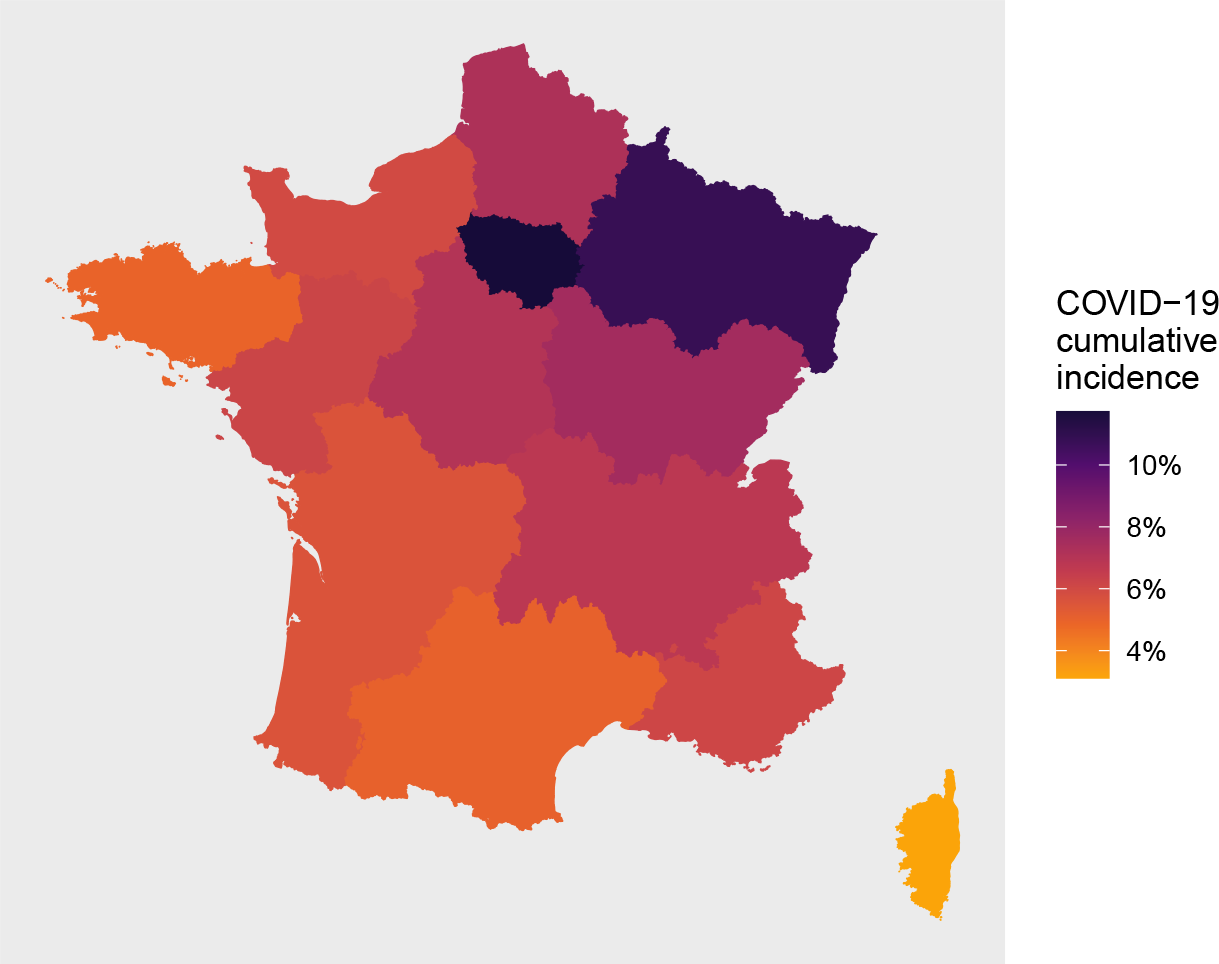
Regional cumulative incidence of COVID-19 after the first wave in metropolitan France

Age-specific cumulative incidences, IHR and IFR are presented in Table 1. The two groups with the highest cumulative incidences were 30-39 year-old persons (13.6%, 95% CI from 12.7 to 14.4%) and 40-49 year-old persons (13.5%, 95% CI from 12.8 to 14.1%). IHR and IFR varied strongly with age, peaking respectively at 33.7% (95% CI from 26.2 to 13.3%) and 21.6% (95% CI from 16.8 to 27.8%) in persons older than 80 years.

**Table 1.**
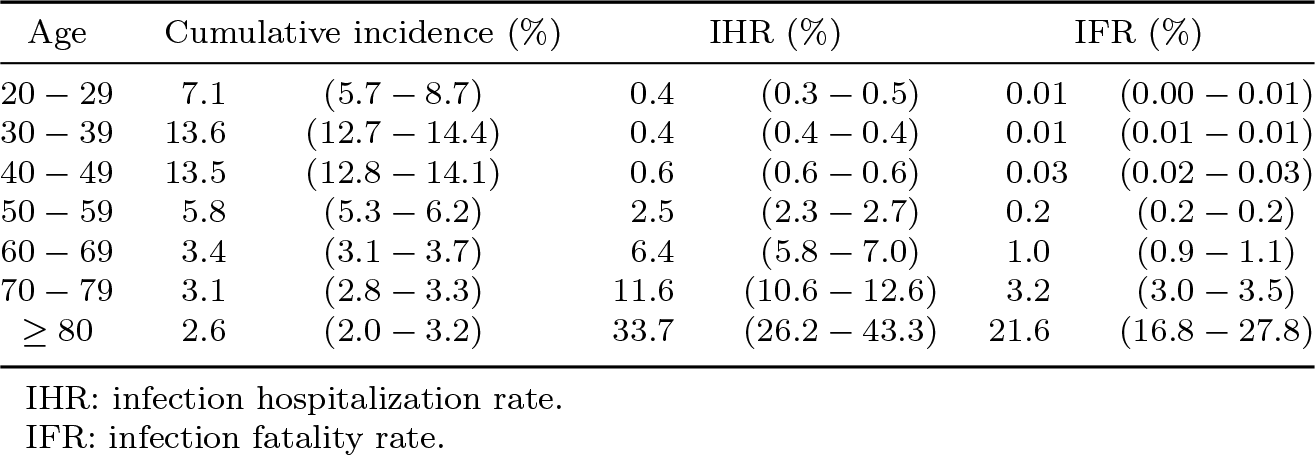
Cumulative incidence and infection-outcome rates depending on age (mean estimates and 95% credible intervals)

### 2.3 Distribution of ELISA and cut-off based test characteristics

Observed and inferred ELISA ODR distributions are displayed in Figure 2. Among infected individuals, the proportion of non-responders was estimated to be 14.5% (95% CI, 11.2% to 18.1%). These distributions imply an AUC of 92.6% (95% CI, 90.0% to 94.9%). Comparing the two manufacturer cut-offs (0.8 and 1.1 ODR) in terms of Younden’s J statistic favors the 0.8 cut-off, with a mean absolute difference of 5.0% (95% CI, 3.9% to 6.2%). Estimated sensitivities, specificities and Younden’s J statistics are displayed in Table 2.

**Table 2.** Characteristics of the serological test depending on the cut-off.

|  | Cut-off: 0.8 | Cut-off: 1.1 |
| --- | --- | --- |
| Sensitivity (%) | 81.3 (77.0 – 85.7) | 76.0 (71.4 – 80.7) |
| Specificity (%) | 99.7 (99.7 – 99.7) | 100 (100 – 100) |
| J statistic* | 81.0 (76.7 – 85.4) | 76.0 (71.4 – 80.6) |

\*Younden's J statistic

**Fig. 2.**
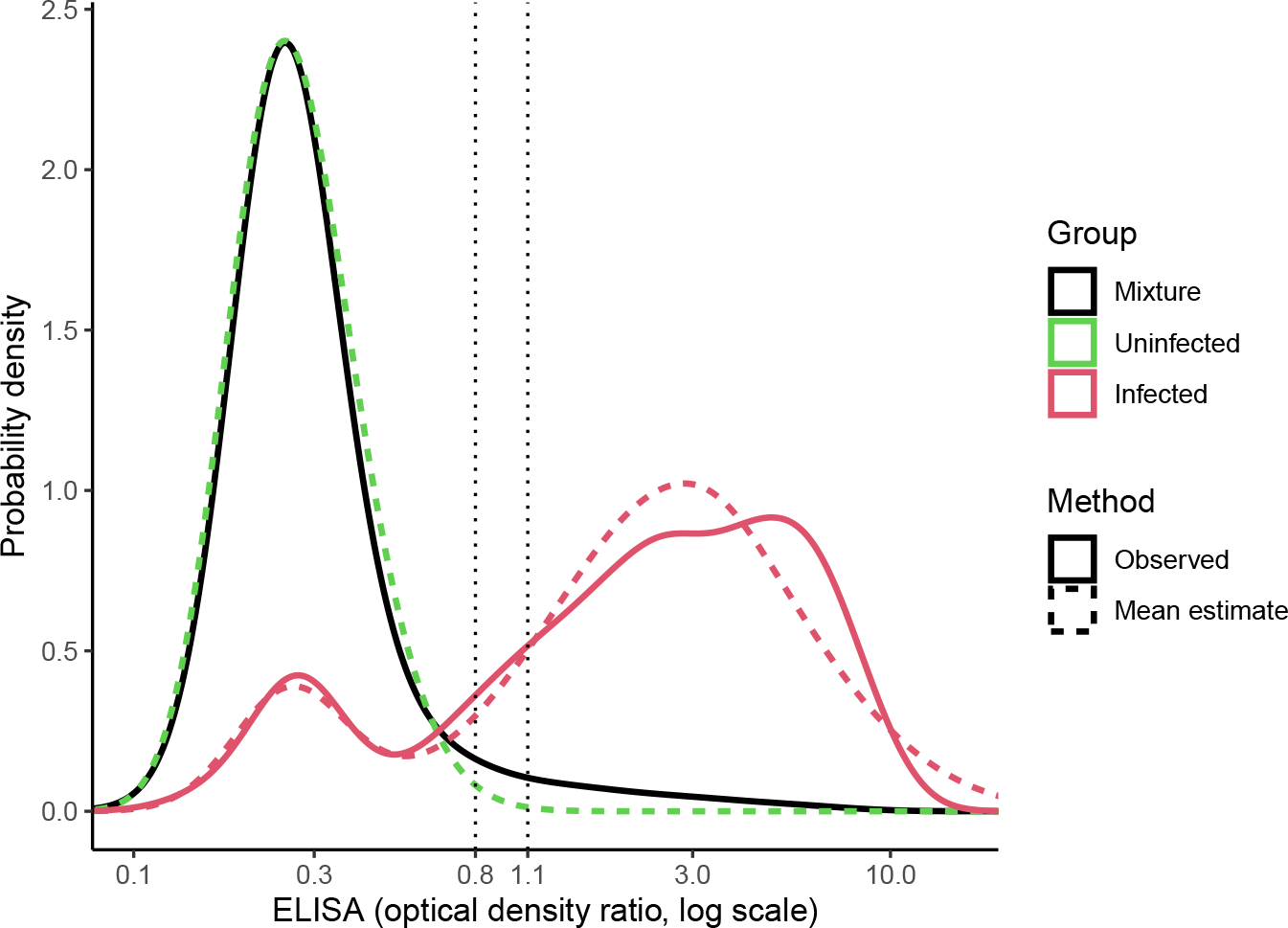
Observed and inferred ELISA distributions

### 2.4 COVID-19 retrospective diagnosis: an illustrative example

Figure 3 illustrates how the probability of having been infected is related to the ODR in two regions and three age groups representing the range of contextual cumulative incidences. Given a negative ELISA test, the infection probability could be as high as 61.9% (95% CI, 54.0 to 68.1%), corresponding to an ELISA result of 0.8 for a person of 40-49 years living in Île-de-France (region with the highest cumulative incidence).

**Fig. 3.**
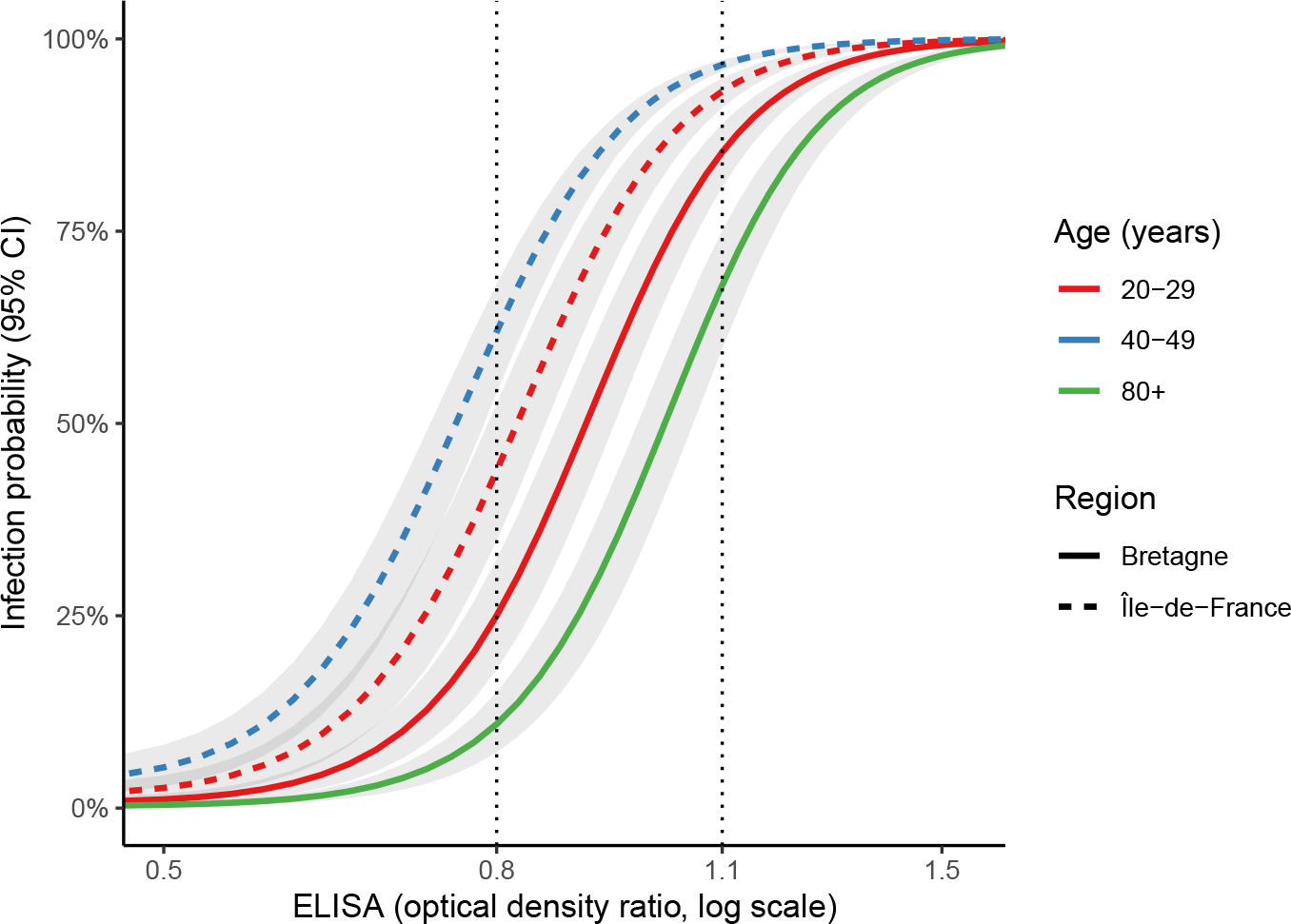
Influence of age, region and ELISA ODR on the probability of infection

Conversely, a positive ELISA was compatible with a probability of infection as low as 68.0% (95% CI, 59.7 to 75.1%), corresponding to an ELISA result of 1.1 for a person older than 80 years living in Bretagne (region with the lowest cumulative incidence, excepting Corsica).

When considering a person of 60-69 years living in Nouvelle-Aquitaine (for example), an intermediate ELISA value could lead to infection probabilities as diverse as 15.7% (95% CI, 11.5 to 19.8%) if the ODR was 0.8 and 76.5% (95% CI, 71.8 to 80.6%) if the ODR was 1.1.

An exhaustive interactive table returning infection probability given age, region and ELISA is provided in Supplementary Table 4 (Supplementary media file).

## 3 Discussion

### 3.1 Interpretation

Implementing a mixture model allowed for the estimation of continuous distributions of ELISA in infected individuals as well as in uninfected ones, even if no data of known uninfected individuals were available. By showing how an ELISA result from a tri-chotomous test (negative, intermediate or positive) could encompass various infection predictions due to different contexts and ELISA ODR, this study helps to explain surprising serological results during the first wave of COVID-19 (such as individuals having had specific COVID-19 symptoms but negative ELISA results). These situations can in fact be compatible with a high probability of infection in areas and age groups with high cumulative incidences if the ODR is not too low.

Another finding from this cohort and model is the size of the “non-responding” sub-population, estimated to be about 14.5% of the infected group. However, like any clustering method, this part of the mixture model provided no further characterization of this population beyond the parameters of *P* (ELISA_NR_), namely an expected value and a standard deviation. In the current work, a major benefit of having modeled this sub-group was the quality of the model’s fit to the data.

This study estimated COVID-19 cumulative incidence at 7.6% after the first wave in France, close to the one estimated in seroprevalence studies (about 5% in the whole country, and 10% in the most affected areas) with independent data or different methods [8–10, 22]. Likewise, the highest cumulative incidence between 30 and 49 years was in line with the higher seroprevalence previously reported in these age groups [8]. Infection-hospitalization and infection-fatality rates rose at exponential paces with age in adults, in a similar magnitude of those previously reported [22–26].

### 3.2 Limitations and assumptions

Several modeling hypotheses were made. First, the distribution of ELISA values in infected individuals did not take age into account, whether directly or through the proportion of non-responders. Similarly, the decrease of antibody levels with time was not modeled, based on the slow waning of anti-spike 1 IgG reported after a natural SARS-CoV-2 infection and since time between infection and testing could not exceed nine months in the current study [5].

The survey spread over 6 months (from early May to early November). Nevertheless, serological tests were analyzed regardless of the sampling date. This strategy was justified by two assumptions: that anti-spike IgG would remain stable, and that new infections were unlikely to occur over this time lapse. The weekly incidence between the first and the second wave was indeed very low, as reflected by a low number of hospitalizations. Supplementary Fig. 1 displays the timing of sampling in comparison with the timing of hospitalizations in France during this period.

Another limitation was due to identification issues, which probably arose from a low cumulative incidence (resulting in a unimodal observed distribution for the mixture, as displayed in Figure 3), enforcing the fitting aside of infected individual’s ELISA distribution (as described in section 4.6). As a consequence, the uncertainty in cumulative incidence was under-estimated (only mean estimates of ELISA distribution in infected persons having been used at this step). This uncertainty was however restored when computing sensitivity, AUC and infection probability (except for its part of uncertainty due to cumulative incidence). This sequential approach (known as the plug-in principle) had a second drawback, as a potential spectrum bias (if present) could not be taken into account (ELISA distribution being only estimated from the known infected individuals).

Lastly, inevitably arbitrary choices for likelihood functions were made. A skew normal distribution was specified for ELISA in the uninfected group, and a mixture of two normal distributions handled the skewness of the overall ELISA distribution in the infected group. These specifications are similar to those of other published mixture models for SARS-CoV-2 serological data, yet adding some extra flexibility at the cost of a few more parameters [17].

## 4 Methods

### 4.1 Serological data

The SAPRIS-SERO survey serological data, previously described, were used in the present study [9, 10, 27]. Based on the SAPRIS cohort (including three general population based adult cohorts), randomly selected participants over 18 year old with regular access to the internet and living in France were invited to take a dried-blood spot by themselves. Samples were sent to a virology laboratory (Unité des virus émergents, Marseille, France) for serological analysis using a commercial ELISA test (Euroimmun, Lübeck, Germany) detecting anti-SARS-CoV-2 IgG directed against the S1 domain of the spike protein of the virus. The results of ELISA assays performed using dried-blood spot samples demonstrated a 98.1 to 100% sensitivity and a 99.3 to 100% specificity with conventional serum assays as a standard [28, 29]. A maximum of one test per participant was performed. Participants reporting a positive RT-PCR test were considered infected. Ethical approval and written or electronic informed consent were obtained from each participant before enrollment in the original cohort. The SAPRIS-SERO study was approved by the Sud-Mediterranée III ethics committee (approval 20.04.22.74247) and electronic informed consent was obtained from all participants for dried blood spot testing. The study was registered (#NCT04392388).

### 4.2 Hospital and demographic data

The French population structure by 10-year age class and administrative region came from the Insee 2020 census (Institut national de la statistique et des études économiques) [30]. The data about COVID-19-related hospitalizations before the 1st of July 2020, by 10-year age class or by region (not both simultaneously) were obtained from SIVIC, the exhaustive national inpatient surveillance system used during the pandemic [31]. The data about general population mortality attributed to COVID-19 before the 1st of July 2020 were obtained from the CépiDc (Centre d’épidémiologie sur les causes médicales de décès) [32].

### 4.3 Model

The statistical analysis was carried out within a Bayesian framework, estimating a posterior joint distribution of parameters and possibly including non-uniform prior distributions. In the rest of this section, prior distributions are not always explicitly written. If so, the distribution in question is uniform.

Serological results, originally expressed as optical density ratios (ODR), were modeled after a logarithmic transformation to be compatible with the use of unbounded probability functions. In the following, *P* (ELISA) refers to the distribution of the log-ODR of serological assays, *I* refers to the set of age classes (10-year groups, from 20 to 90 years, with persons older than 90 included in the 80-89 years group) and *J* is the set of French administrative regions. The distribution of ELISA in the undetermined group (unknown infection status) in an age class *i ∈ I* and a region *j ∈ J*, which is *P* (ELISA|*i, j*), was modeled as a mixture of the distributions *P* (ELISA_+_)and *P* (ELISA_*−*_) of ELISA in infected and uninfected individuals, respectively, with a proportion *p*_*i,j*_ of persons having been infected (which is the cumulative incidence given *i* and *j*):

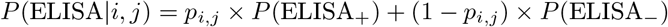

In uninfected individuals, ELISA results were modeled with a skew-normal distribution. The distribution of ELISA in infected individuals was itself a mixture of two normal distributions, the one of responders (ELISA_R_) and the one of non-responders (ELISA_NR_), with *p*_NR_ as the proportion of non-responders (this two-component distribution allowing for some skewness). A prior beta distribution for *p*_NR_ was specified to imply a prior 95% credible interval (95% CI) ranging from 1 to 40% (and thus covering the 5 to 24% estimates previously reported):

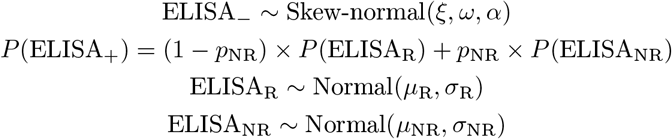

Cumulative incidence on the logit scale, for an age class *i ∈ I* and a region *j ∈ J*, was the sum of a regional intercept and a log-odds ratio of age, without interaction:

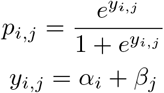

A weakly informative normal prior distribution was specified for age log-odds ratios (*β*_*i*_), with mean 0 and standard deviation 1.

### 4.4 Post-stratification and infection-outcome rates

To correct for a selection bias consisting in differences in age and geographical structures between the French population and the SAPRIS-SERO cohort, age-specific cumulative incidences were reconstructed by post-stratification from *p*_*i,j*_ terms, considering the population sizes pop_*i,j*_ of the different groups:

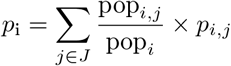

Similarly, metropolitan France’s cumulative incidence was obtained from *p*_*i*_ terms:

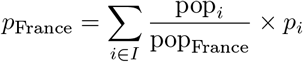

Overall and age-specific IHR (infection-hospitalization rates) and IFR (infection-fatality rates) were then computed as ratios of the number of hospitalizations or deaths to the number of infected persons.

### 4.5 Infection probability given serological results and context

The probability *p*_*x,i,j*_ of having been infected given an ELISA value *x*, an age group *i* and a region *j* was computed using Bayes’ rule. With *P* (*x* | infected) and *P* (*x*| uninfected) being the probability densities of the ELISA value *x* in the infected and uninfected groups, respectively,

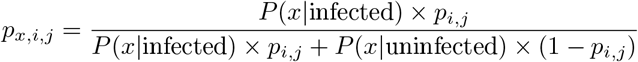

### 4.6 Algorithm and software

The data management used the R software version 4.2.3 and the modeling was done with the Stan software (R package cmdstanr version 0.5.3), which implements Hamiltonian Monte Carlo [33, 34]. The Monte Carlo sampling consisted in 6 chains of 2 000 iterations each (including 1 000 warm up iterations). Trace plots, 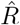 statistics and effective Monte Carlo sample sizes provided by Stan were used to assess convergence. The model’s code (in Stan) is provided in Supplementary Code 1.

Due to identification issues when fitting the mixture model, the distribution of ELISA values in infected individuals was estimated aside (319 persons with positive RT-PCR, see below). Mean parameters’ estimates of this distribution were then introduced as data in the main model, following the plug-in principle [35, 36]. When computing the quantities derived from this distribution in the main model, namely sensitivity, AUC (area under the receiver operating characteristic curve) and infection probabilities, uncertainty was restored by drawing from posterior distributions of the first model’s parameters (more precisely, in normal approximations of these posterior distributions) at each MCMC iteration.

#### Supplementary information

The Supplementary information file attached with this article contains:

- Tables of the participants’ age and region (Supplementary tables 1 and 2)
- A table of exhaustive cumulative incidence estimates along with the number of samples per region (Supplementary table 3)
- A figure of weekly COVID-19 related hospitalizations and serological tests in
- SAPRIS-SERO (Supplementary figure 1)
- The Stan code for the model (Supplementary code 1)
- The list of the SAPRIS-SERO study groups’ members (Supplementary note 1)
- An interactive table returning the probability of infection given ELISA, age and region (Supplementary table 4, in the Supplementary media file)

## Supporting information

Supplementary media

## Data Availability

All data produced in the present study can be made available upon reasonable request to, after a consultation with the steering committee of the SAPRIS-SERO study.

## Acknowledgments

The authors warmly thank all the volunteers of the Constances, E3N–E4N, and NutriNet-Santé cohorts. We thank the staff of the Constances, E3N–E4N and NutriNet-Santé cohorts that have worked with dedication and engagement to collect and manage the data used for this study and to ensure continuing communication with the cohort participants. We thank the CEPH-Biobank staff for their adaptability and the quality of their work. We thank all the members of the SAPRIS and SAPRIS-SERO study groups (Supplementary Note 1).

## Declarations

### Funding

#### SAPRIS-SERO study

ANR (Agence Nationale de la Recherche, #ANR-10-COHO-06), Fondation pour la Recherche Médicale (#20RR052-00), Inserm (Institut National de la Santé et de la Recherche Médicale, #C20-26). The sponsor and funders facilitated data acquisition but did not participate in the study design, analysis, interpretation or drafting. Cohorts funding:

The CONSTANCES Cohort Study is supported by the Caisse Nationale d’Assurance Maladie (CNAM), the French Ministry of Health, the Ministry of Research, the Institut national de la santé et de la recherche médicale. CONSTANCES benefits from a grant from the French National Research Agency [grant number ANR-11-INBS-0002] and is also partly funded by MSD, AstraZeneca, Lundbeck and L’Oreal. The E3N-E4N cohort is supported by the following institutions: Ministère de l’Enseignement Supérieur, de la Recherche et de l’Innovation, INSERM, University Paris-Saclay, Gustave Roussy, the MGEN, and the French League Against Cancer. The NutriNet-Santé study is supported by the following public institutions: Ministère de la Santé, Santé Publique France, Institut National de la Santé et de la Recherche Médicale (INSERM), Institut National de la Recherche Agronomique (INRAE), Conservatoire National des Arts et Métiers (CNAM) and Sorbonne Paris Nord.

The CEPH-Biobank is supported by the " Ministère de l’Enseignement Supérieur, de la Recherche et de l’Innovation ".

### Ethics approval and consent to participate

Ethical approval and written or electronic informed consent were obtained from each participant before enrolment in the original cohort. The SAPRIS-SERO study was approved by the Sud-Mediterranée III ethics committee (approval #20.04.22.74247) and electronic informed consent was obtained from all participants for DBS testing. The study was registered (#NCT04392388).

### Consent for publication

Participants can not be identified on the basis of this article

### Availability of data and materials

In regards to data availability, data from the study are protected under the protection of health data regulation set by the French National Commission on Informatics and Liberty (Commission Nationale de l’Informatique et des Libertés, CNIL). The data can be made available upon reasonable request to, after a consultation with the steering committee of the SAPRIS-SERO study. The French law forbids us to provide free access to SAPRIS-SERO data; access could however be given by the steering committee after legal verification of the use of the data. Please, feel free to come back to us should you have any additional question.

### Code availability

The Stan code for the model is provided as Supplementary Code 1 Authors’ contributions: F.C., N.L. and B.G. conceived and designed the study. B.G. implemented the model and wrote the manuscript. F.C., N.L., X.L., G.S., M.T. and J.-F. D. contributed to data collection. All the authors reviewed and edited the manuscript.

