## Supplementary information for "Refining COVID-19 retrospective diagnosis with continuous serological tests: a Bayesian mixture model"

### Table of contents

**Supplementary Table 1: Region of the participants** 1

**Supplementary Table 2: Age of the participants** 2

**Supplementary Table 3: Exhaustive regional cumulative incidence estimates** 2

**Supplementary Figure 1: Weekly COVID-19 related hospitalizations in France and serological tests in SAPRIS-SERO** 3

**Supplementary Code 1: Stan code** 3

**Supplementary Note 1: the SAPRIS-SERO study group** 9

### Supplementary Table 1: Region of the participants

| Region | N |
|---|---|
| Hauts-de-France | 4892 |
| Normandie | 2769 |
| Île-de-France | 15444 |
| Grand Est | 7146 |
| Bretagne | 5846 |
| Pays de la Loire | 4598 |
| Centre-Val de Loire | 5010 |
| Bourgogne-Franche-Comté | 2757 |
| Nouvelle-Aquitaine | 10579 |
| Auvergne-Rhône-Alpes | 9526 |
| Occitanie | 8130 |
| Provence-Alpes-Côte d'Azur | 5015 |

| Region | N |
|---|---|
| Corse | 85 |

### Supplementary Table 2: Age of the participants

| Age | N |
|---|---|
| 20-29 | 1574 |
| 30-39 | 8808 |
| 40-49 | 14681 |
| 50-59 | 14814 |
| 60-69 | 16642 |
| 70-79 | 22170 |
| 80+ | 3108 |

### Supplementary Table 3: Exhaustive regional cumulative incidence estimates

| Region | Mean (%) | q2.5 (%) | q97.5 (%) | N |
|---|---|---|---|---|
| Île-de-France | 11.7 | 11.0 | 12.4 | 15444 |
| Grand Est | 10.8 | 9.9 | 11.7 | 7146 |
| Bourgogne-Franche-Comté | 7.5 | 6.3 | 8.8 | 2757 |
| Hauts-de-France | 7.2 | 6.3 | 8.2 | 4892 |
| Centre-Val de Loire | 7.0 | 6.2 | 8.0 | 5010 |
| Auvergne-Rhône-Alpes | 6.8 | 6.1 | 7.4 | 9526 |
| Pays de la Loire | 6.2 | 5.3 | 7.1 | 4598 |
| Provence-Alpes-Côte d'Azur | 6.1 | 5.3 | 7.1 | 5015 |
| Normandie | 5.9 | 4.8 | 7.1 | 2769 |
| Nouvelle-Aquitaine | 5.6 | 5.0 | 6.1 | 10579 |
| Occitanie | 5.0 | 4.4 | 5.6 | 8130 |
| Bretagne | 4.9 | 4.2 | 5.6 | 5846 |
| Corse | 3.1 | 0.1 | 10.1 | 85 |

### Supplementary Figure 1: Weekly COVID-19 related hospitalizations in France and serological tests in SAPRIS-SERO

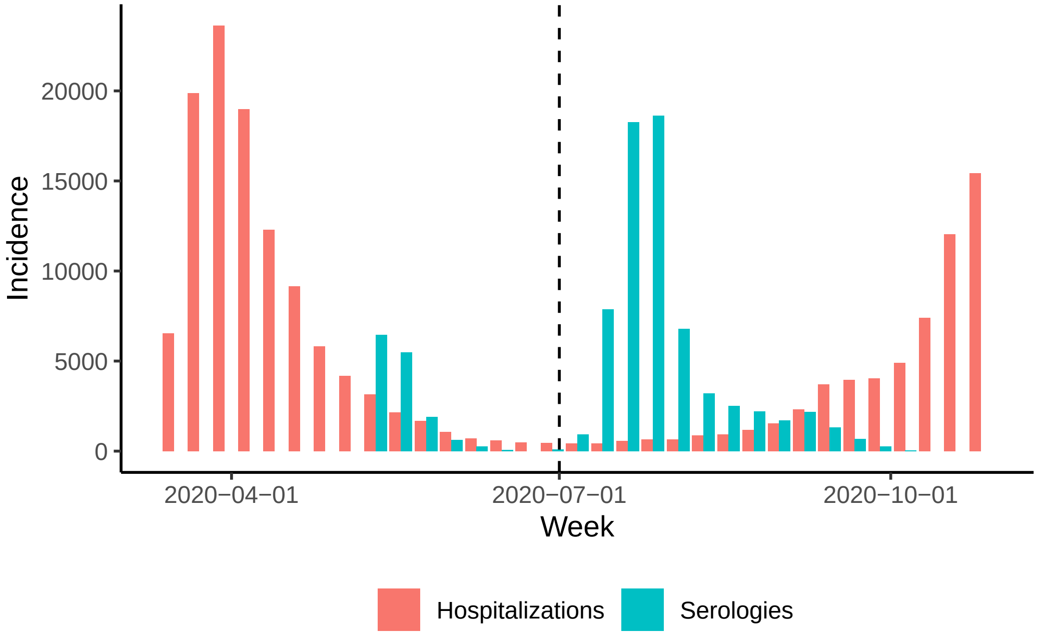


### Supplementary Code 1: Stan code

This first code bloc corresponds to the model of ELISA ODR (optical density ratio) in known infected individuals.

```
data {
  int<lower=0> N_pos;
  vector[N_pos] elisa_pos;
}


parameters {
  ordered[2] mu;
  vector<lower=0>[2] sigma;

  real<lower=0, upper=1> prop_non_resp;
}
```

```
model {

  prop_non_resp ~ beta(1.41, 8.6); // implies a prior 95\% CI ranging from 1 to 40\%

  for (n in 1:N_pos) {
    target += log_mix(
      prop_non_resp,
      normal_lpdf(elisa_pos[n] | mu[1], sigma[1]),
      normal_lpdf(elisa_pos[n] | mu[2], sigma[2])
    );
  }

}
```

This second bloc corresponds to the main model (using the former's mean estimates).

```
data {

  real prop_non_resp;
  real mu_resp;
  real mu_non_resp;
  real<lower=0> sigma_resp;
  real<lower=0> sigma_non_resp;

  real sigma_prop_non_resp;
  real sigma_mu_resp;
  real sigma_mu_non_resp;
  real<lower=0> sigma_sigma_resp;
  real<lower=0> sigma_sigma_non_resp;

  int<lower=0> N_mixt;
  vector[N_mixt] elisa_mixt;

  int<lower=0> N_region;
  array[N_mixt] int region;

  int<lower=0> N_age;
  array[N_mixt] int age;

  matrix<lower=0, upper=1>[N_age, N_region] mat_age_times_region;
  matrix<lower=0, upper=1>[N_region, N_age] mat_region_times_age;
```

```
  vector<lower=0, upper=1>[N_region] prop_region;
  vector<lower=0, upper=1>[N_age] prop_age;
  vector<lower=0, upper=1>[N_age] prop_hospit_age;
  real<lower=0> hospit_tot;
  vector<lower=0, upper=1>[N_age] prop_deces_age;
  real<lower=0> deces_tot;
  real<lower=0> pop_tot;

  int<lower=0> length_grid_sero;
  vector[length_grid_sero] grid_sero;

}


parameters {

  vector[N_age] log_or_age;
  vector[N_region] intercept_region;

  real mu_neg;
  real<lower=0> sigma_neg;
  real alpha_neg;

}


transformed parameters {

  matrix<lower=0, upper=1>[N_age, N_region] incid_age_region;
  for (i in 1:N_age) {
    for (j in 1:N_region) {
      incid_age_region[i, j] = inv_logit(intercept_region[j] + log_or_age[i]);
    }
  }

}


model {

  log_or_age ~ normal(0, 1);
```

```
  for (n in 1:N_mixt) {
    target += log_mix(
      incid_age_region[age[n], region[n]],
      log_mix(
        prop_non_resp,
        normal_lpdf(elisa_mixt[n] | mu_non_resp, sigma_non_resp),
        normal_lpdf(elisa_mixt[n] | mu_resp, sigma_resp)
      ),
      skew_normal_lpdf(elisa_mixt[n] | mu_neg, sigma_neg, alpha_neg)
    );
  }

}


generated quantities {

  // post-stratified incidences

  vector<lower=0, upper=1>[N_age] incid_age;
  for (i in 1:N_age) {
    incid_age[i] = incid_age_region[i, ] * mat_region_times_age[, i];
  }

  real<lower=0, upper=1> incid_fr_via_age = incid_age' * prop_age;

  vector<lower=0, upper=1>[N_region] incid_region;
  for (i in 1:N_region) {
    incid_region[i] = mat_age_times_region[, i]' * incid_age_region[, i];
  }

  real<lower=0, upper=1> incid_fr_via_region = incid_region' * prop_region;

  // infection-outcome rates

  vector<lower=0>[N_age] ihr_age;
  for (i in 1:N_age) {
    ihr_age[i] = prop_hospit_age[i] / incid_age[i];
  }

  vector<lower=0>[N_age] ifr_age;
```

```
for (i in 1:N_age) {
  ifr_age[i] = prop_deces_age[i] / incid_age[i];
}

real<lower=0> ihr = hospit_tot / pop_tot / incid_fr_via_age;

real<lower=0> ifr = deces_tot / pop_tot / incid_fr_via_age;

// drawing from normal approximations of the first model's posteriors

real sample_mu_resp = normal_rng(mu_resp, sigma_mu_resp);
real sample_sigma_resp = normal_rng(sigma_resp, sigma_sigma_resp);
real sample_mu_non_resp = normal_rng(mu_non_resp, sigma_mu_non_resp);
real sample_sigma_non_resp = normal_rng(sigma_non_resp, sigma_sigma_non_resp);
real sample_prop_non_resp = normal_rng(prop_non_resp, sigma_prop_non_resp);

// diagnostic performance of the serological test

real<lower=0, upper=1> sens08 = 1 -
    normal_cdf(log(.8) | sample_mu_non_resp, sample_sigma_non_resp) *
    sample_prop_non_resp -
    normal_cdf(log(.8) | sample_mu_resp, sample_sigma_resp) *
    (1 - sample_prop_non_resp)
  ;

real<lower=0, upper=1> sens11 = 1 -
  normal_cdf(log(1.1) | sample_mu_non_resp, sample_sigma_non_resp) *
  sample_prop_non_resp -
  normal_cdf(log(1.1) | sample_mu_resp, sample_sigma_resp) *
  (1 - sample_prop_non_resp)
;

real<lower=0, upper=1> spe08 =
skew_normal_cdf(log(.8) | mu_neg, sigma_neg, alpha_neg);
real<lower=0, upper=1> spe11 =
skew_normal_cdf(log(1.1) | mu_neg, sigma_neg, alpha_neg);

real<lower=0, upper=1> younden08 = sens08 + spe08 - 1;
real<lower=0, upper=1> younden11 = sens11 + spe11 - 1;

vector<lower=0, upper=1>[length_grid_sero] sens;
```

```
for (i in 1:length_grid_sero) {
  sens[i] = 1 -
    normal_cdf(grid_sero[i] | sample_mu_non_resp, sample_sigma_non_resp) *
    sample_prop_non_resp -
    normal_cdf(grid_sero[i] | sample_mu_resp, sample_sigma_resp) *
    (1 - sample_prop_non_resp)
  ;
}

vector<lower=0, upper=1>[length_grid_sero] c1_spe;
for (i in 1:length_grid_sero) {
  c1_spe[i] = 1 - skew_normal_cdf(grid_sero[i] | mu_neg, sigma_neg, alpha_neg);
}

real<lower=0, upper=1> AUC = (sens[1] + 1)/2 * (1 - c1_spe[1]);
for (i in 2:length_grid_sero) {
  AUC += (sens[i] + sens[i-1])/2 * (c1_spe[i-1] - c1_spe[i]);
}
AUC += sens[length_grid_sero]/2 * c1_spe[length_grid_sero];

// infection probability (given serology, age and region)

array[N_age, N_region, length_grid_sero] real pred_infection_age_region;
for (i in 1:N_age) {
  for (j in 1:N_region) {
    for (k in 1:length_grid_sero) {
    pred_infection_age_region[i, j, k] =
      exp(log_mix(
        sample_prop_non_resp,
        normal_lpdf(grid_sero[k] | sample_mu_non_resp, sample_sigma_non_resp),
        normal_lpdf(grid_sero[k] | sample_mu_resp, sample_sigma_resp)
    )) * incid_age_region[i, j] /
      (
        exp(log_mix(
          sample_prop_non_resp,
          normal_lpdf(grid_sero[k] | sample_mu_non_resp, sample_sigma_non_resp),
          normal_lpdf(grid_sero[k] | sample_mu_resp, sample_sigma_resp)
    )) * incid_age_region[i, j] +
        exp(skew_normal_lpdf(grid_sero[k] | mu_neg, sigma_neg, alpha_neg)) *
        (1 - incid_age_region[i, j])
      );
```

```
      }
    }
  }

}
```

### Supplementary Note 1: the SAPRIS-SERO study group

Fabrice Carrat, Pierre-Yves Ancel, Marie-Aline Charles, Gianluca Severi, Mathilde Touvier, Marie Zins (SAPRIS-SERO investigators).
Sofiane Kab, Adeline Renuy, Stephane Le-Got, Celine Ribet, Mireille Pellicer, Emmanuel Wiernik, Marcel Goldberg, Marie Zins (Constances cohort).
Fanny Artaud, Pascale Gerbouin-Rérolle, Mélody Enguix, Camille Laplanche, Roselyn Gomes-Rima, Lyan Hoang, Emmanuelle Correia, Alpha Amadou Barry, Nadège Senina, Gianluca Severi (E3N-E4N cohort).
Julien Allegre, Fabien Szabo de Edelenyi, Nathalie Druesne-Pecollo, Younes Esseddik, Serge Hercberg, Mélanie Deschasaux, Mathilde Touvier (NutriNet-Santé cohort).
Marie-Aline Charles, Pierre-Yves Ancel, Valérie Benhammou, Anass Ritmi, Laetitia Marchand, Cecile Zaros, Elodie Lordmi, Adriana Candea, Sophie de Visme, Thierry Simeon, Xavier Thierry, Bertrand Geay, Marie-Noelle Dufourg, Karen Milcent (Epipage2 and Elfe child cohorts).
Delphine Rahib, Nathalie Lydie (Santé Publique France).
Clovis Lusivika-Nzinga, Gregory Pannetier, Nathanael Lapidus, Isabelle Goderel, Céline Dorival, Jérôme Nicol, Olivier Robineau, Fabrice Carrat (IPLESP – methodology and coordinating data center).
Cindy Lai, Liza Belhadji, Hélène Esperou, Sandrine Couffin-Cadiergues (Inserm).
Jean-Marie Gagliolo (Institut de Santé Publique).
Hélène Blanché, Jean-Marc Sébaoun, Jean-Christophe Beaudoin, Laetitia Gressin, Valérie Morel, Ouissam Ouili, Jean-François Deleuze (CEPH-Biobank).
Laetitia Ninove, Stéphane Priet, Paola Mariela Saba Villarroel, Toscane Fourié, Souand Mohamed Ali, Abdenour Amroun, Morgan Seston, Nazli Ayhan, Boris Pastorino, Xavier de Lamballerie (Unité des Virus Emergents).
